# Longitudinal analysis of antibody trajectories and humoral responses to a third dose of mRNA vaccines against SARS-CoV-2 in patients with a history of anti-CD20 therapy (RituxiVac 2.0)

**DOI:** 10.1101/2021.11.19.21266572

**Authors:** Daniel Sidler, Alexander Born, Simeon Schietzel, Michael P. Horn, Daniel Aeberli, Jennifer Amsler, Burkhard Möller, Linet M. Njue, Cesare Medri, Anne Angelillo-Scherrer, Luca Borradori, S. Morteza Seyed Jafari, Susanne Radonjic-Hoesli, Andrew Chan, Robert Hoepner, Ulrike Bacher, Laila-Yasmin Mani, Joseena Mariam Iype, Franziska Suter-Riniker, Cornelia Staehelin, Michael Nagler, Cédric Hirzel, Britta Maurer, Matthias B. Moor

## Abstract

**Background:** Morbidity and mortality of COVID-19 is increased in patients with B-cell depleting therapies, the majority of which also show compromised vaccination-induced immune responses. Herein, we report on the trajectories of anti-SARS-CoV-2 antibodies in patients from the original RituxiVac study compared to healthy volunteers and investigate the immunogenicity of a third vaccination in previously non-responding patients.

**Methods:** A follow-up evaluation was performed in volunteers and patients from the RituxiVac Study (NCT04877496), which investigated the humoral and cell-mediated immune response after SARS-CoV-2 mRNA vaccination in patients with a history with anti-CD20 depleting therapies (rituximab or ocrelizumab). The current population included 33 patients and 26 healthy volunteers with initial humoral vaccine response and 32 non-responding patients. Primary outcome was anti-SARS-CoV-2 antibody trajectories in vaccine responders 4.3 months (median; interquartile range [IQR]: 3.6 – 4.8 months) after first evaluation and humoral responses after a third vaccine dose in previous non-responders. Antibody decay rates were compared using analysis of covariance in linear regression.

**Results:** In patients with detectable anti-spike IgG antibodies after two-dose vaccination, circulating anti-spike IgG persisted in 88% (29/33) of patients 5.6 months after the second vaccination (median; IQR: 5.1-6.7) compared to 92% (24/26) of healthy volunteers 6.8 months after the second dose (IQR: 6.0-7.1) (p=0.7). Antibody decay rates were comparable between patients and controls with −0.54 signal/cut-off (s/c) units per month (IQR −0.72 to −0.45) and −0.60 s/c units per month (IQR: −0.88 to −0.44), p=0.70. Two-dose responders with loss of circulating antibodies at follow-up (n=4/33, 12%) had lower initial antibody concentrations (p<0.01). Biomarkers for immunocompetence, including CD3, CD4 or CD19 cell count at baseline did not predict anti-spike IgG persistence. In two-dose non-responders, a third dose of mRNA vaccine against SARS-CoV-2 elicited humoral response with detectable anti-spike IgG antibodies in 19% (6/32) participants. No clinical parameters nor biomarkers of immunocompetence predicted humoral response after a third vaccine dose.

**Conclusion:** The present study reveals comparable antibody reduction rates between patients with CD20-depleting treatment history and healthy volunteers, but inefficient humoral responses to a third dose of SARS-CoV-2 mRNA vaccines in the majority of two-dose non-responders. There is a need for individually tailored vaccination strategies in immunocompromised patients that could be stratified by B cell counts and initial level of antibody titers. (Funded by Bern University Hospital, ClinicalTrials.gov number, NCT04877496)

## Introduction

COVID-19 has a deleterious effect in many patients, including those treated with immunosuppressive drugs. Additionally, increasing evidence supports the finding that mRNA-based vaccines elicit inferior responses in immunocompromised patients, especially in those with anti-CD20 therapies.^1,2^ In the initial RituxiVac Study, we showed that in a mixed population of patients with kidney transplant, autoimmune disease or cancer, only 49% of patients produce a humoral immune response after mRNA vaccination against SARS-CoV-2.^3^ Our recent meta-analysis revealed similar results by several studies, with some differences depending on the types of patient populations or cellular immune assays used.^4^

Recent publications support the notion that patients with a weak humoral immune responses benefit from a third vaccine dose. Indeed, in approximately 30% of solid-organ transplant recipients seroconversion occurred upon a third or fourth SARS-CoV-2 vaccination. ^5–7^ Individual case reports and one larger preprint article revealed increasing humoral and cellular immune responses after a third dose of SARS-CoV-2 vaccination in two-dose non-responders with a history of CD20 treatment.^8–10^ At present, clinical outcome data are lacking to determine whether fully vaccinated but seronegative patients are at least partially protected from severe COVID-19. To add more complexity, even in healthy populations the time frame of protection from severe COVID-19 by SARS-CoV-2 vaccines is under debate,^11^ but much more so in the immunocompromised.^12^ Additionally, it remains to be determined whether antibody trajectories of immunocompromised patients differ from those of healthy individuals.

This study represents a follow-up of patients with treatment history of anti-CD20 therapy from our initial RituxiVac cohort, in which both humoral and cell-mediated immune responses to SARS-COV-2 mRNA vaccines had been assessed in an investigator-initiated, single-center, case-control study (RituxiVac 1.0).^3^ We herein present the 6-months follow-up data on antibody trajectories in initial vaccination responders comparing B-cell depleted patients with healthy volunteers as well as data on humoral immune responses after a third dose of mRNA SARS-CoV-2 vaccines in two-dose non-responders.

## Methods

### Study design

The RituxiVac Study was an investigator-initiated, single center, open-label, case-control study conducted at the Departments of Nephrology and Hypertension, Rheumatology and Immunology, Hematology, Neurology, and Dermatology of the University Hospital in Bern, Switzerland. The study design was previously reported.^3^ In brief, COVID-19-naïve patients with a treatment history of anti-CD20 drugs (rituximab or ocrelizumab) and completion of a two-dose regimen of SARS-CoV-2 mRNA vaccination ≥4 weeks earlier were enrolled between April 26 and June 30, 2021. Treatment data and indication for anti-CD20 therapies were collected, and age, sex and immunosuppressive co-medication were further analyzed. Official vaccination reports or self-reported vaccination dates and types were recorded. Further, unmatched healthy volunteers were enrolled at least four weeks after completion of their two-dose mRNA vaccination. Patients and healthy volunteers aged less than 18 years or with prior SARS-CoV-2 infection were not eligible. All study participants were tested for the presence of anti-nucleocapsid antibodies, and those with positive results were excluded from the analysis.

All participants initially received either BNT162b2 mRNA Covid-19 vaccine (BioNTech/Pfizer, Comirnaty^®^) or mRNA-1273 vaccine (COVID-19 VACCINE Moderna^®^) as issued by the Swiss national COVID-19 vaccination program. Starting July 21^st^ 2021 as per the guidelines of the Swiss Federal Vaccination Commission and independently of the present study, immunocompromised patients who were humoral non-responders after two vaccinations were invited to receive a third dose of BNT162b2 mRNA Covid-19 vaccine or mRNA-1273 vaccine. All RituxiVac participants and two additional patients were contacted by phone or during regular visits and invited to participate in the present follow-up study. This included an assessment either 4 weeks after the third vaccination for initial non-responders, or 6 months (± 2 months) after the second vaccination in initial responders.

This study was supported by Bern University Hospital. The funder had no influence on the design or conduct of this study and was not involved in data collection or analysis, in the writing of the manuscript, or in the decision to submit for publication. The study was registered on clinicaltrials.gov (Identifier: NCT04877496) and was performed in accordance with the principles of the Declaration of Helsinki. The study was approved by the Ethics Committee of the Canton of Bern, Switzerland. All participants provided written informed consent before inclusion. The authors assume responsibility for the accuracy and completeness of the data and analyses, as well as for the fidelity of the study and this report to the protocol.

### Study procedures

#### Baseline data collection

Study nurses and physicians completed a 20-item questionnaire for the follow-up study visit. Dates and types of administered vaccines (BNT162b2 mRNA Covid-19 vaccine or mRNA-1273 vaccine) were obtained from the existing study database, and additional data were retrieved from the official vaccination records when available or were self-reported by participants.

#### Blood collection and measurement of anti-SARS-CoV-2 S1-IgG and NC-IgG

For the detection of SARS-CoV-2 specific antibodies, IgG antibodies against SARS-CoV-2 S1 protein were measured by a commercial ELISA from Euroimmun AG, Lübeck, Germany as previously reported^13^. Samples were diluted 1:100, and 100 μL of diluted samples, prediluted positive or negative controls and a prediluted calibrator were added for 1 hour at 37°C. After washing three times, 100μl of HRP-labelled secondary anti-human IgG was added for 30 minutes at 37°C, followed by three additional washes. Next, 100 μL of a TMB solution was added for 20 minutes. The reaction was interrupted with 100 μL of 0.5M H_2_SO_4_, and the results were obtained by measurement at OD450-620 nm. Antibody results were expressed as ratio (OD_sample_/OD_calibrator_). All samples with a ratio > 1.1 were considered as positive following the manufacturer’s instructions. To allow an exclusion of the participants with previous COVID-19, an anti-nucleocapsid ECLIA test was performed using a Cobas 8000 analyzer (Roche Diagnostics, Rotkreuz, Switzerland).^14^ The cut-off was calculated based on the calibrator measurements, and a cut-off index s/c ≥ 1.0 was considered positive with adherence to the manufacturer’s instructions.

### Outcomes

The primary endpoints were the proportion of patients with a history of anti-CD20 therapies showing a persisting humoral response against SARS-CoV-2 spike protein 6 months after completion of SARS-CoV-2 vaccination, in comparison to immunocompetent controls, and the *de novo* detection of a humoral response against SARS-CoV-2 spike protein in two-dose non-responders that received a third dose of SARS-CoV-2 mRNA vaccines. Humoral responses were defined as anti-SARS-CoV-2 S1 ≥ 1.1 (Index).^15^

Pre-specified secondary endpoints were the rate of decline in anti-S1 IgG concentrations in patients with anti-CD20 therapy and healthy volunteers, and the effects of initial anti-S1 concentration after a two-dose vaccination regimen, demographic data, time since last treatment and cumulative dose of B-cell depleting agents, treatment indication or blood markers of immunocompetence on humoral responses to SARS-CoV-2 mRNA-based vaccines.

### Statistical analysis

For this follow-up study, the initial RituxiVac study population was eligible without further power analysis or pre-screening procedures. Statistical analyses were conducted using R software version 4.0.4.^16^ Pearson’s Chi squared test was used for comparison of categorical variables. Wilcoxon rank sum test or t-test were used for comparison of continuous variables as indicated. Linear regression analyses were performed using the lm function in R. Statistical significance was assumed at a two-tailed p<0.05. P values and 95% confidence intervals are not adjusted for multiple testing.

## Results

### Demographic and clinical characteristics of the participants

For the RituxiVac 2.0 Study, 65 patients, including 33 two-dose humoral responders and 32 non-responders, and 26 healthy controls, were assessed at a follow-up visit between September 7th and November 10^th^ 2021, reflecting a follow-up rate of 63% for patients and 81% for volunteers (Supplementary Figure 1). Patients with de novo anti-nucleocapsid antibodies at the follow-up visit were excluded (n= 4 for Patients, n= 1 for Volunteers). Anti-CD20 drugs included rituximab and ocrelizumab and were prescribed for autoimmune disease in 45 cases (69%), for malignancy in 5 cases (8%), and for ABO-incompatible kidney transplantation in 15 cases (23%). Demographic details, treatment history and vaccination data are presented in Supplementary Table 1. Immunosuppressive co-medication was present in 41 patients (63%) and included corticosteroids in 32 cases (49%), calcineurin inhibitors in 17 cases (26%), antimetabolites in 21 cases (32%), methotrexate in 2 cases (3%), cytotoxic chemotherapy in 1 case (1%), or other immunosuppressive drugs in 3 cases (5%). No healthy volunteer was treated with immunosuppressive drugs.

### Follow-up assessment of antibody trajectories in humoral responders to two-dose SARS-CoV-2 mRNA vaccination

Serum samples of two-dose regimen humoral responders were obtained a median of 6.8 months (IQR: 6.0-7.1) after the second vaccination and 4.6 months (4.4-5.0) after the initial serological assessment in volunteers versus 5.6 months (5.1-6.7) after vaccination and 3.9 months (3.5-4.0) after the first serological assessment in patients (Table 1). We recorded a median spike S1 IgG level of 4.2 (3.1-5.8) Index s/c in volunteers and 3.7 (1.7-5.7) in the two-dose regimen responders with a history of anti-CD20 treatment (p=0.42) (Figure 1A, Table 1). Anti-S1 IgG concentrations above the manufacturer’s cut-off were present in 92% of volunteers and 88% of patients at follow-up. In patients with a decline of anti-S1 IgG below detection cut-off, the initial concentration of anti-S1 was lower with 2.26 s/c (1.98-2.43) versus 7.14 s/c (4.89-8.55) in those with persisting antibodies (p<0.01). Moreover, patients with persisting antibodies tended to be younger at study enrollment and had lower rate of immunosuppressive co-medication. Parameters of immunocompetence such as CD4, CD19 counts or total IgM were not associated with antibody persistence, neither was treatment history of rituximab (time since last dose and cumulative dose). In an analysis of covariance using linear regression models, anti-S1 IgG concentrations at follow-up depended on initial anti-S1 concentration (p<0.001) irrespective of anti-CD20 treatment status (p=0.84), indicating a similar rate of anti-S antibody clearance in both patients and healthy volunteers.

**Table 1:** Bivariate analyses of anti-SARS-CoV-2 IgG persistence and third dose humoral response for sets of clinical or laboratory variables important for immune competence in patients with a history of anti-CD20 therapy. V1, first study visit. Positive indicates anti-SARS-CoV-2 IgG above 1.1 (s/c ratio) at V2 visit. Data are presented as median (interquartile range) or counts (frequency of group). Statistical analyses were performed using Pearson’s Chi-squared test or Wilcoxon rank sum test.

|  | Anti-Spike IgG Persistence |  |  | Third Dose Vaccine IgG Response |  |  |
| --- | --- | --- | --- | --- | --- | --- |
|  | anti-S1 IgG negative | anti-S1 IgG positive | p-value | anti-S1 IgG negative | anti-S1 IgG positive | p-value |
|  | n=4 | n=29 |  | n=26 | n=6 |  |
| **Male Sex (%)** | 1 (25%) | 14 (48%) | 0.6 | 14 (54%) | 4 (67%) | 0.7 |
| **Median Age (years)** | 76 (70, 79) | 66 (53, 70) | 0.2 | 66 (54, 72) | 58 (43, 71) | 0.5 |
| **Immunosuppression (%)** | 3 (75%) | 10 (34%) | 0.3 | 22 (85%) | 6 (100%) | 0.6 |
| **Vaccine (Biontech/Pfizer) (%)** | 3 (75%) | 14 (48%) | 0.6 | 19 (73%) | 5 (83%) | >0.9 |
| **Time since last anti-CD20 treatment (years)** | 2.08 (0.90, 3.22) | 1.57 (0.79, 4.44) | 0.8 | 0.80 (0.40, 2.89) | 2.97 (1.01, 3.63) | 0.5 |
| **Cumulative dose anti-CD20 (g)** | 2.25 (2.00, 2.94) | 3.00 (2.00, 5.00) | 0.5 | 2.00 (0.65, 3.00) | 0.65 (0.65, 1.66) | 0.078 |
| **CD4 cells (cells/ul)** | 748 (720, 1,126) | 949 (658, 1,048) | 0.9 | 624 (350, 905) | 638 (427, 814) | >0.9 |
| **CD19 cells (cells/ul)** | 34 (26, 45) | 51 (24, 176) | 0.5 | 3 (0, 21) | 52 (15, 92) | 0.052 |
| **V1 anti-SARS-CoV2 S1 IgG** | 2.26 (1.98, 2.43) | 7.14 (4.89, 8.55) | **0.002** | 0.12 (0.09, 0.15) | 0.16 (0.14, 0.29) | 0.069 |
| **V1 IFNγ release (mU/ml)** | 0.04 (0.03, 0.05) | 0.06 (0.00, 0.32) | >0.9 | 0.000 (0.000, 0.000) | 0.010 (0.000, 0.010) | 0.2 |

**Figure 1:**
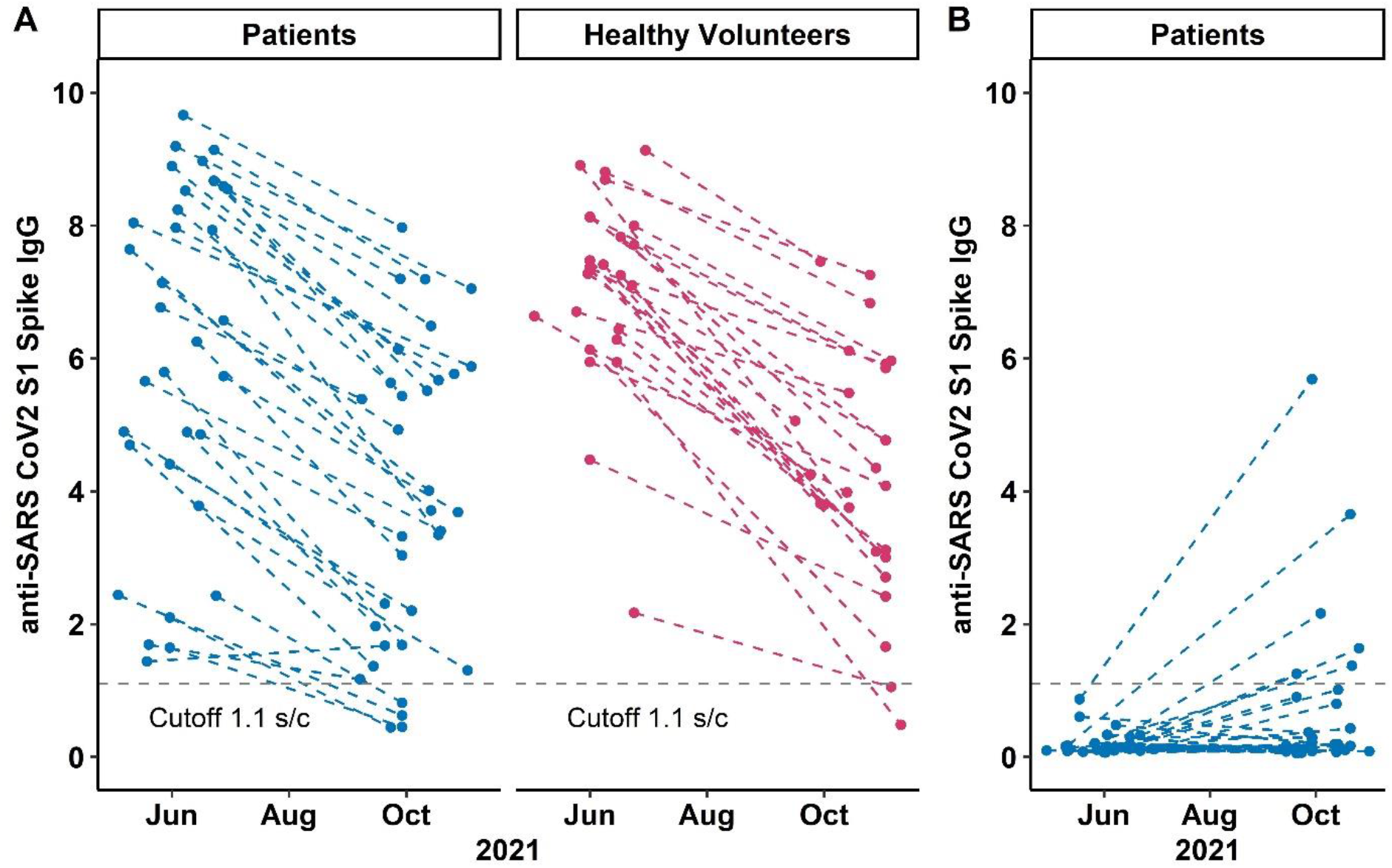
anti-SARS CoV2 S1 Spike IgG levels at study visit 1 and 2 in A) patients and volunteers with two dose humoral response and B) patients with a third dose vaccination. Each point represents one study visit; intraindividual values are connected with dashed lines. The dotted grey line denotes the cut-off anti-SARS-CoV-2 S1-IgG value of 1.1 (signal to cutoff ratio).

### Humoral response to a third mRNA SARS-CoV-2 vaccine dose

Patients without an initial response to a two-dose regimen of SARS-CoV-2 vaccines were given a third dose of mRNA-1273 vaccine (n=8/32) or BNT162b2 mRNA Covid-19 vaccine (n=23/32) after a median period of 5.0 months (4.1-6.0) after their second vaccination dose. The serological assessment revealed an anti-S1 IgG seroconversion in 19% (6/32) of these participants only (Figure 1B). The three-dose responders did not significantly differ from non-responders with regard to demographic, clinical and immunocompetence parameters (Table 1). Third-dose responders tended towards higher CD19 counts at baseline (p=0.052) and and to a lower cumulative dose of anti-CD20 treatments received (p=0.078). Despite the very low patient numbers, we observed a tendency to higher initial anti-S in the three-dose regimen responders with 0.16 (0.14-0.29) s/c compared to non-responders with 0.12 (0.09-0.15) s/c (p=0.069). To summarize, only a small proportion of patients with a history of anti-CD20 treatments underwent a seroconversion after a third SARS-CoV-2 vaccination.

## Discussion

Our longitudinal analysis of 91 participants of the initial investigator-initiated, single center, open-label RituxiVac Study^3^ shows a comparable rate of decline of circulating anti-spike antibodies between patients with anti-CD20 therapies and healthy volunteers at 6 months of follow-up after a two-dose regimen of mRNA SARS-CoV-2 vaccines, with 88% of patients and 92% of volunteers still presenting with detectable antibody levels. Higher initial antibody concentrations were associated with persistence of circulating anti-spike antibodies at follow-up. Moreover, we demonstrate that only a minority of patients with anti-CD20 therapies that were two-dose non-responders developed anti-spike antibodies after a third SARS-CoV-2 vaccination. Again, initial antibody titers but also B cell counts and lower cumulative doses of anti-CD20 treatment tended to associate with seroconversion after a third vaccine. These findings may assist the design of future, more individualised vaccination strategies in this immunocompromised patient population.

The initial RituxiVac study^3^ was finalized shortly before the Swiss Federal Office of Public Health allowed a third dose of SARS-CoV-2 vaccination in immunocompromised patients who did not respond to a two-dose regimen. Therefore, we converted the study design to a longitudinal observational study. Our main finding is thus that after a third dose of SARS-CoV-2 mRNA vaccines only a subset of patients mounts a humoral immune response. Emerging reports by others have found a similar rate of around 25% *de novo* anti-spike seroconversion after a third dose of SARS-CoV-2 vaccines in patients on anti-CD20 therapies that were non-responders after two-dose vaccination regimen.^8–10,17^ Secondly, circulating antibodies decay at similar rates in patients with a history of anti-CD20 therapies compared to healthy volunteers. and the initial titer magnitude is key for the persistence of anti-SARS-CoV2 antibodies at a 6 months period. These data are in agreement with previous reports in the general population^18,19^ and further support the current regimen of the Swiss Federal Office of Public Health to provide all immunosuppressed patients access to a three-dose primary series vaccination regimen.

Since factors such as co-immunosuppression and circulating lymphocyte subpopulations could assist in predicting immune responses to vaccination,^3^ prospective studies should focus on these factors to enable individualized vaccination strategies and to determine optimal timing and number of additional vaccine doses in the immunocompromised.

The present study has some limitations. First, while we observed a decline of circulating antibodies, this is a natural phenomenon because SARS-CoV-2 specific T and B memory cells persist within lymph nodes.^20^ Hence, an assessment of cell-mediated immunity would give additional insight to determine longer-lasting immune responses in anti-CD20 patients as described in a first report.^21^

Next, the present analysis did not include enough longitudinal measurements to allow for an in-depth modeling of the antibody decay dynamics that has been demonstrated in SARS-CoV-2 infection studies ^22,23^ or in selected reports of vaccinated healthy volunteers.^19^ With better understanding of the concentrations of circulating or neutralizing antibodies required for protection of severe disease in immunocompromised patients are understood like in the general population,^24^ prediction models based on neutralizing or total anti-spike antibody clearance rates and circulating peripheral immune cells could guide the administration of future vaccine doses. This is essential, because the heterogeneity of patients with anti-CD20 therapies and their immune response, the differences in their comorbidities, co-immunosuppression and in their environmental exposure to COVID-19 hinder the establishment of a simple generic algorithm for SARS-CoV-2 vaccination in this population. We therefore recommend to closely observe this population and invest in targeted public health strategies for different subsets of immunocompromised patient groups.

## Data Availability

Data are available from the authors on request.

## Acknowledgements

The authors thank all participants of the study and all involved nurses and study nurses including Barbara Strehler, Sabine Hasler, Astrid Zbinden, Mark Wienand, Matthias Gyger, Theres Rath, Sarah Rieder and Ruth Kober for their contributions. Furthermore, the authors wish to thank Monika Hurni, Juliette Schlatter, Olivier Schaerer, Thomas Mormot and Rodoljub Pavlovic for excellent technical assistance.

## Conflict of interest statement

None of the authors declare conflicts of interest.

## Data availability

Data are available from the authors on request.

## Authors’ contributions

MBM and DS conceived the study. MBM, MPH, CH, BM and DS designed the study. MBM, DA, AB, BM, LN, CM, AAS, LB, SR, AC, RH, UB, LYM, BM and DS recruited participants. MBM, SS, AB and DS performed computational chart review. MBM, FRS, MPH, JMI, MN, CH, BM and DS performed analyses. MBM, BM and DS wrote the manuscript with input from all authors. All authors approved the final version of the manuscript.

## Funding

The current study was funded by Bern University Hospital. The funder had no role in design, interpretation of results, manuscript drafting or decision to publish the results.

## Supplement

**Supplementary Table 1:** Baseline characteristics, vaccination history of patients and healthy controls, and anti-CD20 B-cell depletion history of patients in the study

|  | Baseline Characteristics |  | p-value |
| --- | --- | --- | --- |
|  | Patients | Healthy Volunteers |  |
|  | n=65 | n=26 |  |
| **Male Sex (%)** | 33 (51%) | 10 (38%) | 0.3 |
| **Median Age (years)** | 66 (54, 73) | 53 (45, 60) | **0.002** |
| **Vaccine (BioNTech/Pfizer) (%)** | 41 (63%) | 8 (31%) | **0.005** |
| **Time between 2nd Vaccine Dose and V2 Visit (months)** | 5.97 (5.23, 6.87) | 6.75 (5.97, 7.16) | 0.075 |
| **Time between 3rd Vaccine Dose and V2 Visit (months)** | 1.15 (1.00, 1.31) |  |  |
| **Months between V1 and V2 Visit (years)** | 3.90 (3.50, 4.43) | 4.57 (4.35, 5.03) | **0.004** |
| **Cumulative dose anti-CD20 (g)** | 2.30 (1.00, 4.00) |  |  |
| **Time since last dose of anti-CD20 therapy (months)** | 1.46 (0.56, 3.08) |  |  |

**Supplementary Figure 1:**
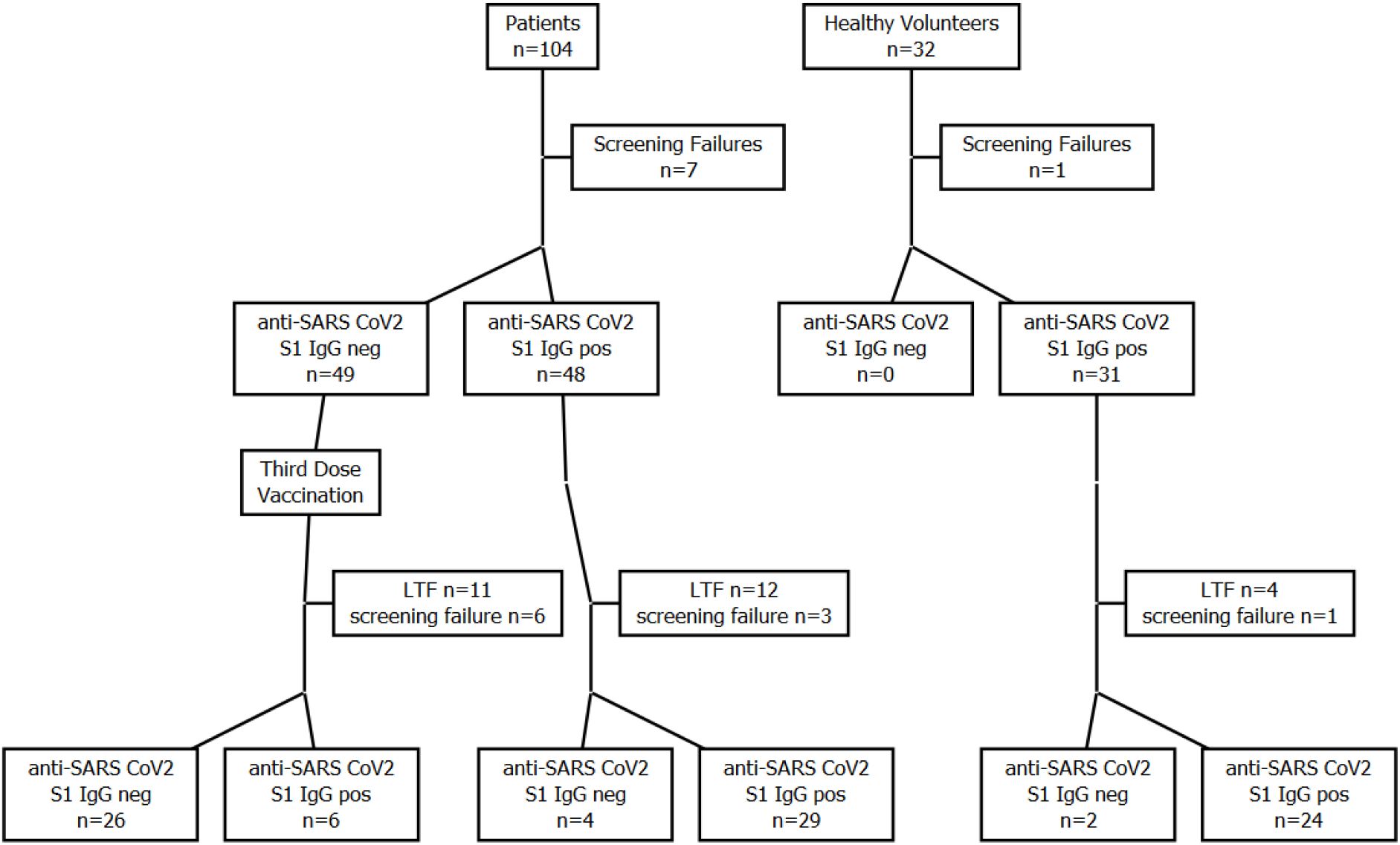
Flowchart of screening and grouping of patients and healthy volunteers in the RituxiVac study. LTF: lost to follow-up, pos=positive, neg=negative

**Supplementary Figure 2:**
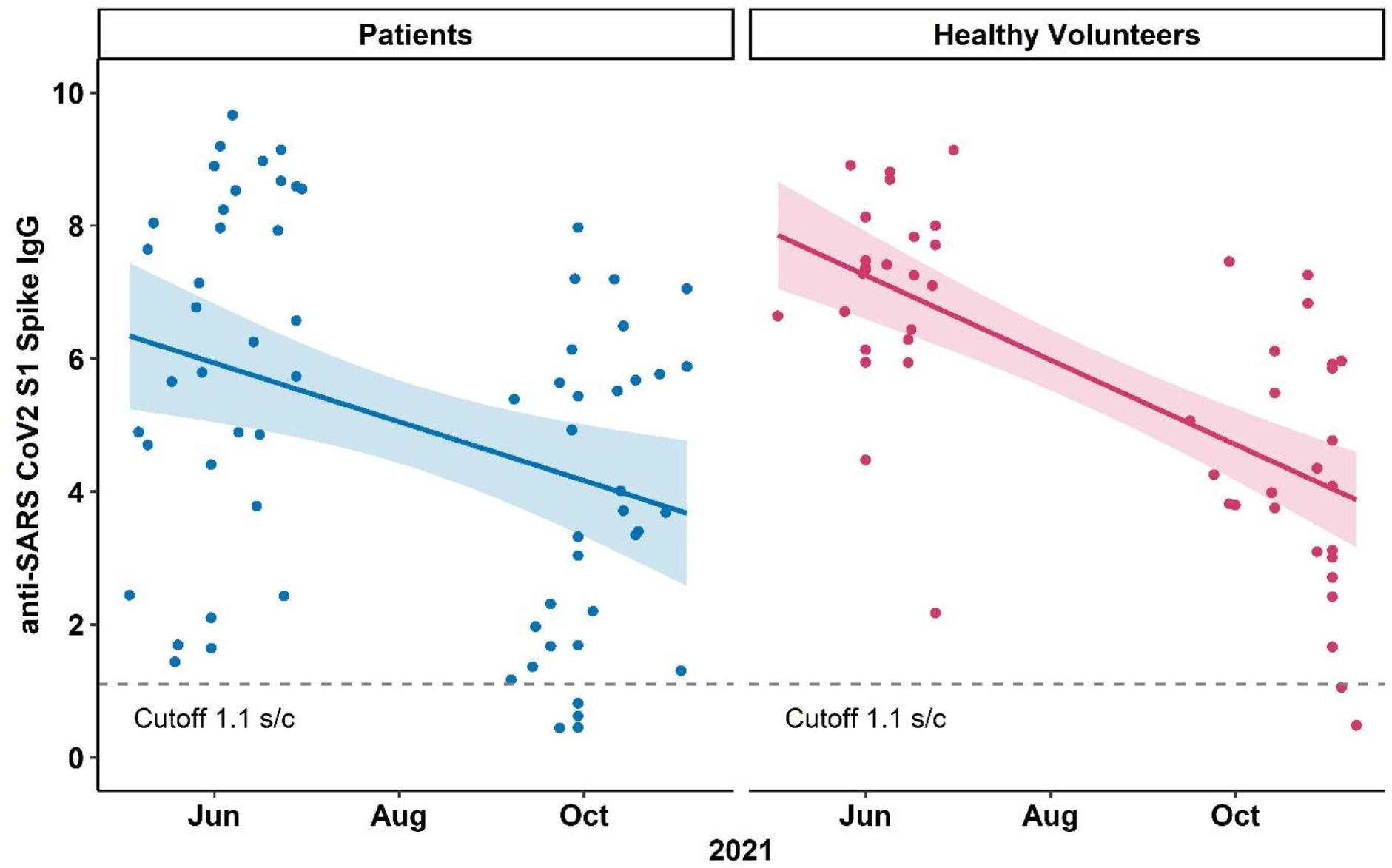
Linear regression between anti-SARS-CoV-2 S1 IgG concentrations and date of study visit. The shaded grey area represents the 95% CI for the regression line. Points denote individual values. Each point represents one patient. The dotted line denotes the cut-off anti-SARS-CoV-2 S1-IgG value of 1.1 (signal to cutoff ratio).

## Notes

### Competing Interest Statement

The authors have declared no competing interest.

### Author Declarations

Kantonale Ethikkommission Bern gave ethical approval for this work.

